# Determinants of first aid knowledge and basic practice among elementary school teachers in Debre Tabor City, Northwest Ethiopia

**DOI:** 10.1101/19005439

**Authors:** Wubet Taklual, Maru Mekie, Chalachew Yenew

**Author notes:** Corresponding Author: Wubet Taklual, Department Public Health, College of Health Sciences, Debre Tabor University, Debre Tabor, Ethiopia, Post office box: 272. Author’s address.

## Abstract

**Background:** Unpremeditated injuries are the leading causes of morbidity and mortality in pediatrics population, especially in low and middle-income countries. Giving immediate help for an injured child is a crucial step for saving the child from further disability and/or death. This study aimed to assess the determinant factors of first aid knowledge and basic practice among elementary school teachers in Debre Tabor, Ethiopia.

**Method:** Institution based cross-sectional study was employed in Debre Tabor City. Single population proportion formula was used for sample size calculation and a total of 216 elementary school teachers were included in study. Simple random sampling technique with proportional allocation was applied for selections of the study participant. Data entry was done by Epi data version 3.1 and the data was exported to SPSS version 21 for analysis. Binary and multivariable logistic regression analysis were performed to identify determinants of knowledge on first aid. Crude and adjusted odds ratios were used to determine the significance and strength of association at 95% confidence interval.

**Result:** Our study revealed that 45.8% of the subjects were knowledgeable on first aid. More than 75% of study participant reported that they have encountered a child who need first aid. Among this 64% of them provide first aid. The multivariable analysis revealed that service year (AOR=3.51, 95%CI: (1.06, 11.59)), educational status (AOR=12.15, 95%CI: (3.17, 46.67)), previous first aid training (AOR=0.43, 95%CI: (0.21, 0.87)) and information about first aid (AOR=0.12, 95%CI ;(0.03, 0.48)) were found to be significantly associated with having knowledge on first aid.

**Conclusion:** School teachers have low knowledge on first aid. Educational status, service year, previous first aid training and information on first aid were the predictor of first aid knowledge. Introducing essential first aid training in the curriculum during teachers’ training shall be considered.

## Introduction

First-aid is commonly defined as the “immediate assistance given at the time of injury or sudden illness by a stander or other lay person before the arrival of expert medical aid”(1). First aid is by no means a replacement for emergency services; it is a vital initial step for providing effective and immediate action that helps to reduce serious injuries and improve the chances of survival. Emergency services’ response time is clearly since the likelihood of an injured person living or dying is dependent on the timeliness of life-saving measures(2).

Injuries are the major causes of morbidity and mortality in the world, especially in middle and low-income countries (3, 4). Globally, in 2013, there were 973 million people sustained injuries of which 77.9% was an unintentional injury that warranted some types of care (4). Injuries among school-age children are common in developing countries, which accounts for 13% of all diseases pattern and disability (5). Unintentional injuries are the leading causes of morbidity and mortality in pediatrics children (5, 6). Childhood DALY (Disability Adjusted Life Year) which attributed to injuries were higher in sub-Saharan African countries (5). In Ethiopia, injuries caused 25 thousand deaths among 0-14-year olds and unintentional injuries accounts the majority. The country longitudinal projection survey suggests that there will be 27,807, and 30,364 death due to injuries by the year 2020 and 2030 respectively (7).

School-age children are more likely to experience unintentional injuries in the school, while they are playing (8). Teachers in the school are the primarily responsible body for keeping the welfare of the pupils and oversee their activities. They are the first contact and responsible person when children face injuries (8). Schools have no nurse or physician who will give first aid in Ethiopia. Pre-hospital school-based emergency medical service (EMS) at school by school personnel is mandatory for saving the pupils from disability and death attributed to injury-related problems (9). Therefore, addressing knowledge and basic practice gaps of a school teacher on first aid is vital, especially at the elementary school level. To the best of researchers’ knowledge, school teachers’ knowledge and basic practice on first aid in Ethiopia, isn’t clearly known. Thus, this study is the first study in Ethiopia which assesses the teachers’ knowledge and basic practice on first aid in elementary schools.

## Methods

### Study design and study population

An institution based cross-sectional study design was employed in Debre Tabor city elementary school teachers from April/2018-June/2018. There are 10 government and private elementary schools in Debre Tabor run by a total of 422 teachers. The source population was all elementary school teachers who are teaching in Debre Tabor city. Whereas, randomly selected elementary school teachers were taken as the study population. A total of 216 elementary school teachers were selected by simple random sampling method among the selected government and private schools proportionally.

### Sample size determination and data collection method

The sample size was performed by using a single population proportion formula. By considering the following assumptions, Za/2=1.96, p=0.4(a study conducted in Addis Ababa among kindergarten teachers (10), d (margin of error) =5%. Moreover, a finite population correction formula was applied since the source population is < 10,000. Thus, the final sample size becomes 216 after adding 10% non-response rate.

The data was collected by using pretested, structured self-administer questionnaire. The questionnaire was prepared in English then translated into Amharic (local language) and then back to English for consistency by the language experts. Information about socio-demographic characteristics of the study participants, first aid knowledge and basic practice were collected by using the self-administered questionnaires. Knowledge on first aid was assessed by using 20 knowledge assessment questionnaires. Whereas, basic first aid practice was assessed by 16 questionnaires. The quality of the data was ensured during data collection, coding, entry, and analysis. In addition, the content validity, face validity, feasibility, sequence, and flows of the questionnaires were checked.

### Data analysis and processing

The data were coded and entered in a template prepared in Epi-data Version 3.1. Data cleaning and editing were performed by SPSS by running frequencies and cross-tabulations. Descriptive statistics were performed by using frequencies and percentages in a table. Binary logistic regression was done to identify the determinant factors of knowledge such as age, sex, types of school, educational status, service year, previous first aid training and information about first aid). Hosmer-Leme show goodness of fit test was used to test the fitness of the model. Variables with a p-value of < 0.2 in the binary logistic regression were included in the multivariable model to single out the independent predictors of knowledge. The statistical significance declared at p<0.05 with 95% confidence level.

### Ethical approval

Ethical clearance was obtained from the Research Review Committee (REC) of the college of health sciences, Debre Tabor University. An official letter was written to South Gondar education department. Written and verbal informed consent was obtained from all respondents who were participated in the study. Autonomy of the participants and the confidentiality of the information were maintained.

## Results

### Socio-demographic characteristics of the study participants

With regards to age distribution, 71 (32.9%) and 29(13.4 %) of the study participants were found in ages of 25-29 and 30-34 years respectively. The sex distribution of the study participants was almost proportional with 107(49.5%) and 109(50.5%) for males and females respectively. More than half (137 (63.4%) of the elementary school teachers were diploma holders. Only a third of the respondents 76 (35.2%) were trained on first aid. With regards to information about first aid, 175 (81%) had heard about first aid. As per participants self-report, health personnel was mentioned as a main source of information by 121 (69.1%) of the participants (see table 1 for the detail)

**Table 1:**
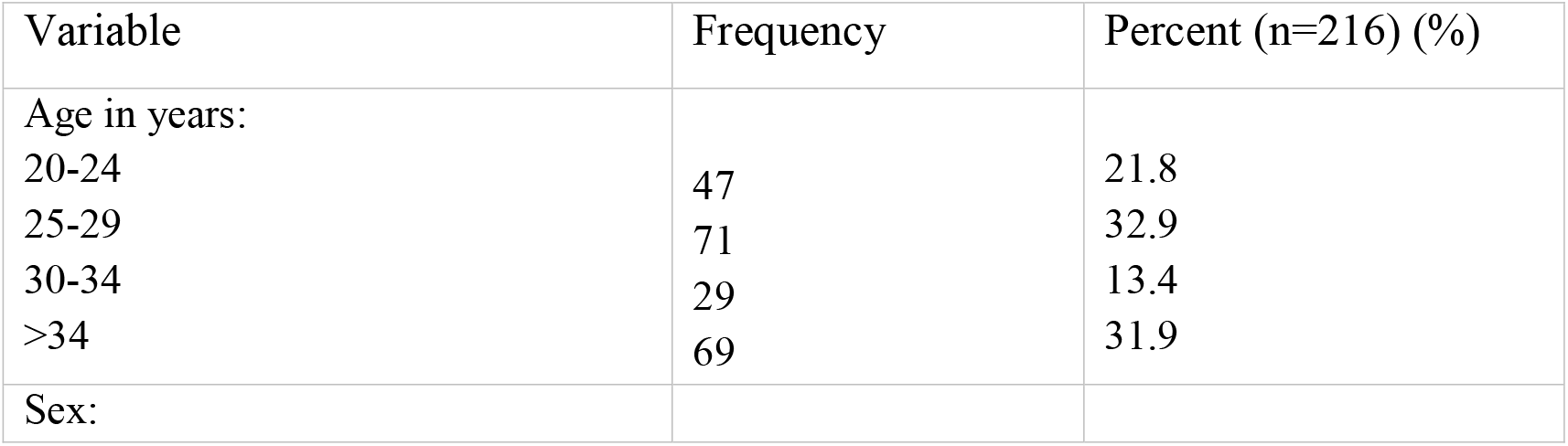
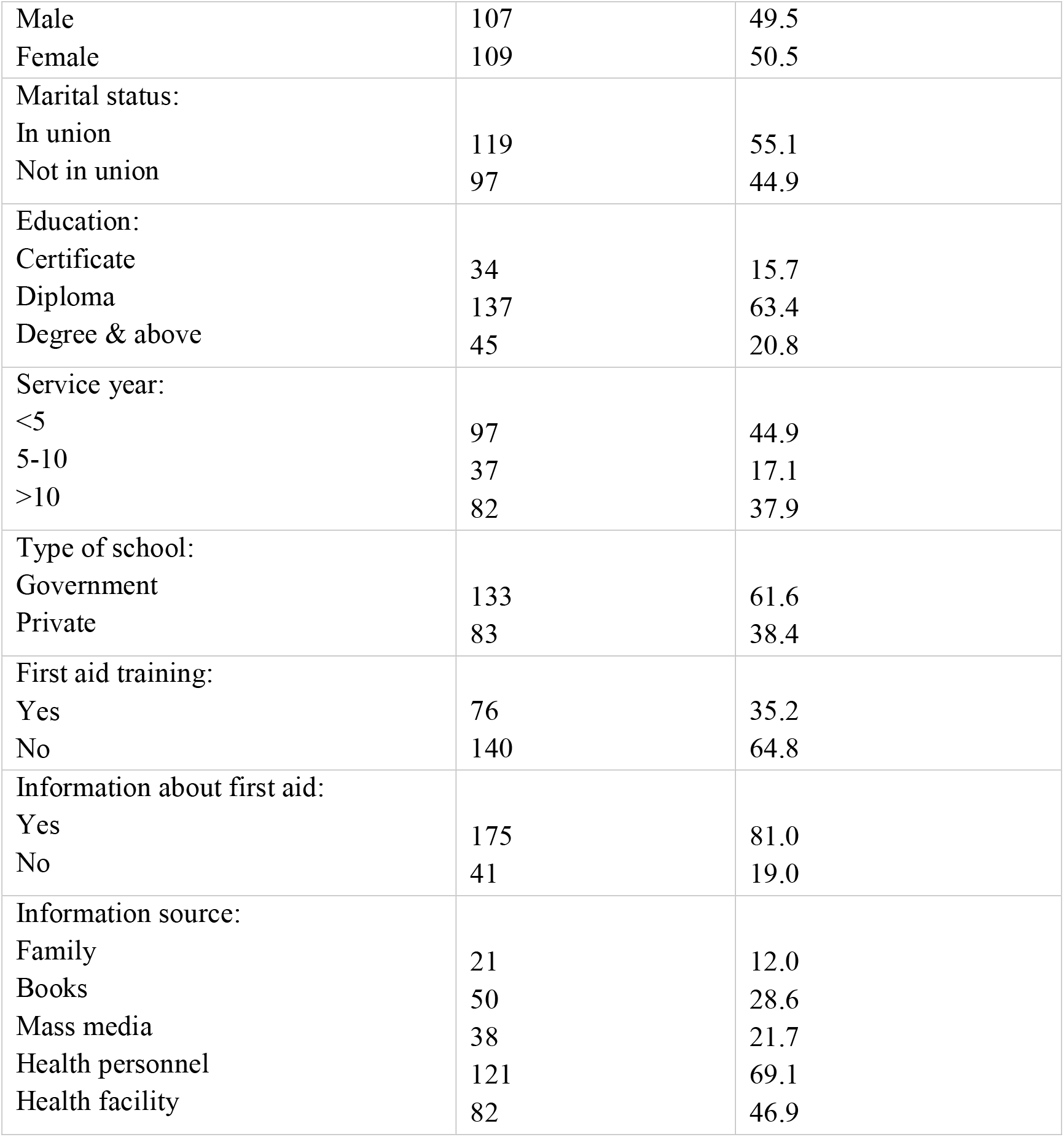
Socio-demographic characteristics of elementary school teachers in Debre Tabor City

### Knowledge of elementary school Teachers on first aid

The knowledge of the study participants was assessed by using 20 knowledge assessment questions. The cases were selected based on the frequency of encounter in the school setting. The questions were yes/no questions which were designed to address emergency cases. Those participants who scored above the mean score of knowledge assessment question were considered as knowledgeable. Whereas, those who scored below the mean score were considered as not knowledgeable in our study. The mean score of knowledge assessment questions was 11.6 with a minimum and maximum score of 0 and 20 points respectively. Of the total study participants, 99 (45.8%) scored above the mean value of knowledge questions which implies more than half of the study participants, 117 (54.2%) had no knowledge about first aid.

Based on the respondents’ point of view, whether the cases need first aid or not. Most of the respondents 190(88%) agreed on needs of first aid in bleeding. As per participants understanding, more than half of the respondents, epilepsy 112(51.9%), choking 129(59.7%), nasal bleeding 123(56.9%), fainting 127(58.8%), swallowed poison 112(51.9%) and breathing difficulty 111(51.4%) needs first aid. Whereas, fracture 83(38.4%), human/ animal bite 63(29.2%), burning 100(46.3%) and neck and back injuries 53(24.5%) were responded by less than half of the participants as they need first aid.

Most of the respondents knew the correct measures against stopping bleeding 186(86.1%) followed by choking147 (68.1%), epilepsy 133(61.6%), epistaxis/nose bleeding 154(71.3%), and breathing difficulty 145(67.1%).

### Practices of Elementary school Teachers on First aid

Table 2 below reveals the practice of elementary school teachers on first aid. About seventy-five percent of teachers face a child who needs first aid, among them 158 (96.9 %) of teachers took an action. Of 158 teachers who took an action, 5 (3.1%), 52 (34.2%), and 101 (64%), calling ambulance, transfer the victim to a nearby health institution, and give an appropriate first aid respectively. Seventy-six (35.2 %) of the study participants faced a child with difficulty of breathing. Of whom the majority encouraged the victim to sit quietly 60 (79 %), breath slowly and deeply 49 (64.5%). With regards to fainting, more than one-third of the respondents 84 (39.4%) had faced a child with fainting. The majority of the respondents 64 (76.2%) reported as they put the victim on flat surface. Epistaxis was another emergency case which was faced by 145 (67.1%) study participants. Of whom 115 (79.3%) help the child by encouraging to sit comfortably with slightly forward position as first aid measure. One hundred four (48.1%) respondents faced body bleeding and most of them were pressed firmly with a clean bandage to stop bleeding 87 (83.7%) (See table: 2).

**Table 2:** Practice of elementary school teachers about first aid in Debre Tabor City

| Variable | Frequency (No) | Percent (%) |
| --- | --- | --- |
| Faced a child within need of first aid |  |  |
| Yes | 163 | 75.5 |
| No | 53 | 24.5 |
| Teacher took an action |  |  |
| Yes | 158 | 96.7 |
| No | 5 | 3.3 |
| Their first actions |  |  |
| Calling ambulance | 5 | 3.1 |
| Transfer to the hospital | 52 | 32.9 |
| Give appropriate first aid | 101 | 64.0 |
| Breathing difficulty |  |  |
| Yes | 76 | 35.2 |
| No | 140 | 64.8 |
| Measure for Breathing difficulty |  |  |
| Call ambulance | 19 | 25.0 |
| Help to sit quietly | 60 | 78.9 |
| Breath slowly/deeply | 49 | 64.5 |
| Contact authority | 35 | 46.1 |
| Child with fainting |  |  |
| Yes | 85 | 39.4 |
| No | 131 | 60.6 |
| Measures for fainting |  |  |
| Call ambulance | 19 | 22.6 |
| Help to sit quietly | 64 | 76.2 |
| Loosen clothes around | 38 | 45.2 |
| Clear airway | 43 | 51.2 |
| offer nothing per mouth | 25 | 29.8 |
| Contact authority | 25 | 29.8 |
| Epistaxis: |  |  |
| Yes | 145 | 67.1 |
| No | 71 | 32.9 |
| Measures for epistaxis: |  |  |
| Call ambulance | 22 | 15.2 |
| Help to Sit slightly forward | 115 | 79.3 |
| Laid on the side by a pillow | 59 | 40.7 |
| Apply pressure on a nostril | 68 | 46.9 |
| Apply ice on the nose | 64 | 44.1 |
| Body Bleeding: |  |  |
| Yes | 104 | 48.1 |
| No | 112 | 51.9 |
| Bleeding management: |  |  |
| Call ambulance | 15 | 14.4 |
| Bandage bleeding site | 87 | 83.7 |
| Elevate bleeding part | 35 | 33.7 |
| Bandage with no pressure with circulation | 69 | 66.3 |
| Cover child with a blanket | 21 | 20.2 |
| Contact school authority | 38 | 36.5 |
| Epilepsy: |  |  |
| Yes | 94 | 43.5 |
| No | 122 | 56.5 |
| measures for Epilepsy mgt: |  |  |
| Call ambulance | 17 | 18.1 |
| help to sleep on the floor | 61 | 64.9 |
| left free epileptic movement | 56 | 59.6 |
| move nearby objects to avoid injury | 59 | 62.8 |
| hold any drink/food per mouth | 50 | 53.2 |
| clear airway and placing the child on the side | 37 | 39.4 |
| report to parents/school authority | 29 | 30.9 |
| Choking: |  |  |
| Yes | 100 | 46.3 |
| No | 116 | 53.7 |
| Management of choking: |  |  |
| Call ambulance | 18 | 18.4 |
| Squeeze the child chest turning back chest | 79 | 80.6 |
| compression until expel | 76 | 77.6 |
| Contact responsible body | 24 | 24.5 |

### Factors affecting knowledge of elementary school Teachers on First aid

Variables with a p-value of < 0.2 in the binary logistic regression were included in the multivariable model to single out the independent predictors of knowledge. Thus, 7 variables were included in the final model. Variables such as service year, previous training on first aid, education status and information about first aid were found to be significantly associated with knowledge in the multivariable model at 95% confidence interval. Table 3 below reveals determinants of knowledge of primary school teachers on first aid. The odds of being knowledgeable on first aid was 3.5 times higher among elementary school teachers who had more than five years of experience compared with counterparts (AOR=3.51, 95%, CI: (1.06, 11.59)). Educational levels of the respondents were also found to affect first aid knowledge. Those degree and above holders were 12 times more likely to be knowledgeable than those under degree levels (AOR= 12.15, 95%, CI, (3.17, 46.67)). Participants who have no previous training are less likely to be knowledgeable on first aid than those who had previous training on first aid (AOR=0.43, 95%CI: (0.21, 0.87)). The odds of being less knowledgeable on first aid found to be higher among participants who have had no information about first aid (AOR=0.12, 95%CI ;(0.03, 0.48))(see table 3 for more detail).

**Table 3:**
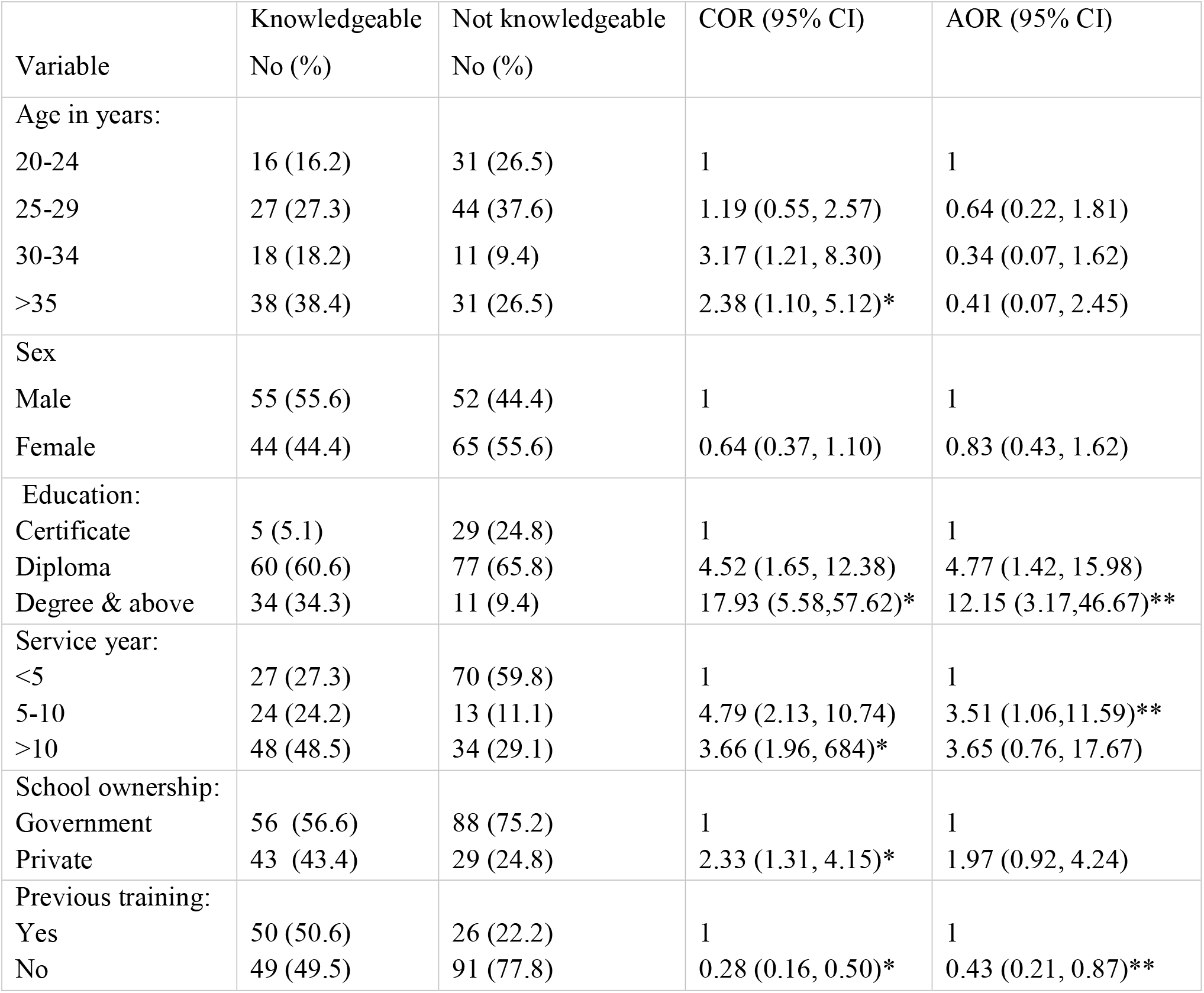
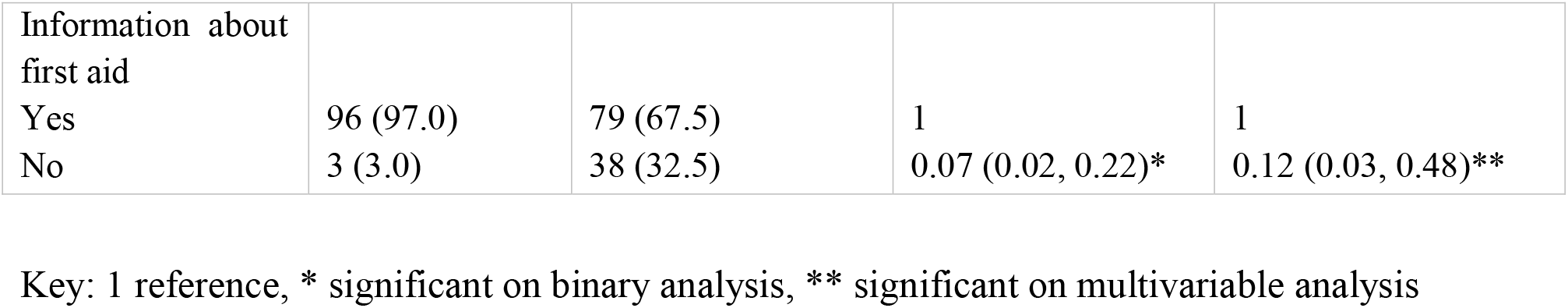
Bivariable and multivariable logistic regression analysis of factors affecting knowledge of elementary school teachers on first aid

## Discussion

Children spend most of their time in school and school teachers are expected to act as a guardian for keeping the welfare of children. Understanding teachers’ level of awareness and basic practice on first aid is moving one step forward for a solution. This study was aimed to assess the knowledge and basic practice of elementary school teachers on first aid and its determinant factors in Debre Tabor City.

The mean score on knowledge assessment question was taken to classify teachers as knowledgeable and not knowledgeable. Our study revealed that more than half of the study participants were less knowledgeable about first aid (54.2%). However, 158 (64%) reported as they provide first aid at least once. Our finding indicated that study participants provide first aid measures without having detail knowledge about the problem. This might be due to a low number of trained staffs (35.2%), the absence of essential first aid training during teacher’s training and poor school health program. Incorporating first aid in the teachers training curricula, strengthening the existing programs, and establish school health program could be a significant measure to fill the knowledge gaps on first aid. The finding of our study is consistent with a study conducted among kindergarten teachers in Addis Ababa, Ethiopia (10), and another study conducted among a sample of elementary school teachers in Kayseri, Turkey (11).

The finding of this study is higher than the study conducted in Assiut city, Egypt among preparatory school teachers which found 97.3% teachers to have poor knowledge score on pretest (12) despite a significant improvement was recorded following a training 75.3% good performance. A similar finding was also reported in a study conducted in Isparta, Turkey among preschool teachers which found only 15.5% participants to have good knowledge on first aid (13). The variations could be explained by variations in sample size in which the aforementioned studies were conducted among 150 and 110 teachers, unlike our study. Variation in measurement could also explain the discrepancy. The finding of our study is also higher than a study conducted in Baghdad, Iraq among primary school teachers with regards to external bleeding and fracture (14). This difference might be due to the variation of the number of trained staffs on first aid, the accessibility of information, and variation in school setup.

With regards to training, one-third of the respondents have taken training on first aid in this study. Our finding is supported by the studies conducted among kindergarten teachers in Addis Ababa which were 32.2% (10), in China 30.8% (15), and Assiut City, Egypt 26.7% (12). Those participants who did not take any training on first aid were found to be less knowledgeable compared with those who had training on first aid (AOR=0.43, 95%CI ;(0.21, 0.87)). This suggests that training is an important strategy to increase the knowledge of school teachers on first aid. The finding of our study is consistent with previous studies conducted in Addis Ababa, Ethiopia among kindergarten school (10), Shanghai, China among preschool teachers (16), and Mangalore school teachers, South India (17). Thus, basic and refreshment training on first aid shall be conducted to increase the knowledge of elementary school teachers on first aid.

With regards to information about first aid, eighty percent of the respondents have information about first aid in this study. The finding is in line with the study in Mangalore, South India (74%) (17). Lower than the finding in Addis Ababa which was (100%). The variation might be explained by variation in the level of awareness between study participants and the setting in which Addis Ababa more urbanized place than Debre Tabor (10). As per participants’ report, health personnel and health facilities were the main sources of information about first aid in our study, unlike previous studies which reported mass media as a primary source of information to first aid (11, 13, 18). The variation might be the difference in coverage of mass media as well as less emphasis of mass media on school health programs and first aid. In our case, those participants who did not have previous information about first aid were less likely to be knowledgeable than counterparts (AOR=0.12, 95%CI ;(0.03, 0.48)). A similar finding was reported in the study in Addis Ababa, Ethiopia in which participants whose source of information were a health professional or health institution were more likely knowledgeable than others (10).

Service year was found to be significantly associated with the level of knowledge about first aid. Consistent with a previous study (10), the multivariable logistic regression analysis revealed that the odds of being knowledgeable were found to be 3.5 times higher among teachers who had more than five years of experience compared to counterparts (AOR=3.51,95%CI; (1.06, 11.59)). This implies that people can learn from experience in addition to the chance of getting training on first aid and emergency medical care through time. The finding of our study revealed that special emphasis shall be given to newly employed teachers since experience plays its role in the acquisition of knowledge on first aid.

Our study showed that there is a significant association between educational status and their knowledge on first aid. The higher the education level, the more likely to be knowledgeable on first aid. Those diploma holder elementary school teachers were found to be more likely to be knowledgeable compared to certificates (AOR=4.77 (1.42, 15.98). Similarly, those who were degree and above were twelve times more likely to be knowledgeable than counterparts (AR=12.15 (3.17, 46.67). A similar finding was reported in the study conducted in Shanghai, China among preschool teachers which found high knowledge about first aid among teachers who had attended college or more level of education(16). This implies that educational status affects the level of understanding.

Regarding the basic practice, seventy five percent of the study participants encountered a child who needs first aid, of whom only 64% (n=158) has provided emergency medical aid, Thirty-three percent and 3.1% take measure by transferring the injured child to the hospital and calling ambulance respectively. The finding of our study showed that a third of elementary school teacher could not provide appropriate emergency first aid at a school level. Thus, special attention shall be given for improving the practice of elementary school teacher on first aid since unintentional injuries are common in a school setting. The finding of our study is consistent with the study in South India which found 36.8% did not provide first aid for those in need and the study in Mysore, India reported 45.4% poor first aid practice among school teachers (19). The finding of our study is not supported by the study in Addis Ababa which found 89.7% of kindergarten teachers provide first aid for children in need (10). Similarly, our finding is inconsistent with other studies in India which reported 72%-84% of school teachers provided first aid (17, 20). The discrepancy may be explained by the difference in availability of first aid room, first aid kit, and emergency drugs in a school setting.

This study shall be seen in consideration of the following limitations. First, the study was conducted in one city which might affect generalizability and representativeness of the finding. Second, the cross-sectional study design was used which may affect causal inference.

## Conclusion and Recommendation

More than half of elementary school teachers did not have knowledge on first aid. Those with an education level of degree and above, having previous training on first aid, availability of information on first aid and work experience of 5 years and more were found to be more knowledgeable than counterparts. Schools should have separate rooms for first aid, first aid kit, and emergency medication. School administrators shall give due emphasis for the training of staffs on first aid and create a link with health institutions. Integrating first aid and school health programs in teachers training curricula shall be considered to reduce childhood DALYs related to an injury.

## Data Availability

The datasets used in this study is available from the corresponding author can be accessible through reasonable request.

## Abbreviations

AOR: Adjusted Odds Ratio
COR: Crude Odds Ratio
SPSS: Statistical Package for Social Science
DALY: Disability-Adjusted Life Year

## Declarations

### Ethical approval and consent

The study was approved by the research ethics committee of the College of Health Sciences, Debre Tabor University. An official letter was submitted to South Gondar zone education department. The purpose of the study was revealed to the study participants and oral consent was obtained from each participant prior to the initiation of the data collection process.

### Consent for publication

Not applicable.

### Competing interest

The authors declare they have no competing interests.

### Funding

No fund was obtained for this particular study.

### Authors’ contribution

WT and MM have contributed in the design, data entry, thesis write-up, manuscript development, and edition. CY has contributed in the data collection process and data cleaning. The final manuscript is reviewed and approved by all authors.

## Acknowledgment

Authors would like to acknowledge the College of Health Science, Debre Tabor University securing the ethical review process. The authors are also want to express the deepest gratitude to supervisors, data collectors, and study participants.

## Notes

### Competing Interest Statement

The authors have declared no competing interest.

### Author Declarations

All relevant ethical guidelines have been followed and any necessary IRB and/or ethics committee approvals have been obtained.

Any clinical trials involved have been registered with an ICMJE-approved registry such as ClinicalTrials.gov and the trial ID is included in the manuscript.

## References

1. Miles S. ASPECTS OF EMERGENCY CARE. British Medical_Journal. 1969;4:485–7.

2. International federation of red cross and red crescent society and european reference center. First aid for a safer future Updated global edition. Geneva International federation of Red cross and Red Crescent society. ; 2010.

3. Global Burden of Disease Pediatrics C, Kyu HH, Pinho C, Wagner JA, Brown JC, Bertozzi-Villa A, et al. Global and National Burden of Diseases and Injuries Among Children and Adolescents Between 1990 and 2013: Findings From the Global Burden of Disease 2013 Study. JAMA pediatrics. 2016;170(3):267–87.

4. Haagsma JA, Graetz N, Bolliger I, Naghavi M, Higashi H, Mullany EC, et al. The global burden of injury: incidence, mortality, disability-adjusted life years and time trends from the Global Burden of Disease study 2013. Injury prevention : journal of the International Society for Child and Adolescent Injury Prevention. 2016;22(1):3–18.

5. Deen J, Vos T, Huttly SRA, Tulloch J. Injuries and noncommunicable diseases: emerging health problems of children in developing countries: World Health Organization.; 1999.

6. Krug EG, Sharma GK, Lozano R. The Global Burden of Injuries. American Journal of Public Health. 2000;90(4):523–6.

7. Li Q, Alonge O, Lawhorn C, Ambaw Y, Kumar S, Jacobs T, et al. Child injuries in Ethiopia: A review of the current situation with projections. PloS one. 2018;13(3):e0194692.

8. Carter YH, Bannon MJ, Jones PW. The role of the teacher in child accident prevention. JOURNAL OF PUBLIC HEALTH MEDICINE. 1994;16(1):23–8.

9. Knight S, Vernon DD, Fines RJ, Nremt- PS., Michael JD. Prehospital Emergency Care for Children at School and Nonschool Locations. Pediatrics 1999;103(6):1–5.

10. Ganfure G, Ameya G, Tamirat A, Lencha B, Bikila D. First aid knowledge, attitude, practice, and associated factors among kindergarten teachers of Lideta sub-city Addis Ababa, Ethiopia. PloS one. 2018;13(3):e0194263.

11. Baser M, Coban S, Tasci S, Sungur G, Bayat M. Evaluating first-aid knowledge and attitudes of a sample of Turkish primary school teachers. Journal of emergency nursing: JEN : official publication of the Emergency Department Nurses Association. 2007;33(5):428–32.

12. Mohamed El magrabi N, ElwardanyAly S, Abdel-Rahim S. Impact of training program regarding first aid knowledge and practices among preparatory schools’ teachers at Assiut City. Journal of Nursing Education and Practice 2017;7(12):89–97.

13. Sonmez Y, Uskun E, Pehlivan A. Knowledge levels of pre-school teachers related with basic first-aid practices, Isparta sample. Turk pediatri arsivi. 2014;49(3):238–46.

14. Al-Robaiaay. YK. Knowledge of Primary School Teachers Regarding First Aid In Baghdad Al-Rusafa. Al – Kindy Col Med J. 2013;9(1):54.

15. Li F, Sheng X, Zhang J, Jiang F, Shen X. Effects of pediatric first aid training on preschool teachers: a longitudinal cohort study in China. BMC Pediatrics. 2014;14(209):1–8.

16. Li F, Jiang F, Jin X, Qiu Y, Shen X. Pediatric first aid knowledge and attitudes among staff in the preschools of Shanghai, China. BMC Pediatrics. 2012;12(121):1–7.

17. Nitin Joseph N, Narayanan T, Zakaria S, Venugopal A, Belayutham N, Aathiya Mihiraa A, et al. Awareness, attitudes and practices of first aid among school teachers in Mangalore, south India. JOURNAL OF PRIMARY HEALTH CARE. 2015 7(4):274–81.

18. Karadag S, Yildirim Z. The Effects of Basic First Aid Education on Teachers’ Knowledge Level: A Pilot Study. International Journal of Caring Sciences 2017;10(2):813–8.

19. Sunil KD, Kulkarni P, Srinivas N, Prakash B, Hugara S, Ashok N. PERCEPTION AND PRACTICES REGARDING FIRST-AID AMONG SCHOOL TEACHERS IN MYSORE. Natl J Community. 2013;4(2):349–52.

20. Masih S, Kumar R, Kumar A. Knowledge and practice of primary school teachers about first aid management of selected minor injuries among children. International Journal of Medicine and Public Health 2014;4(4):458–62.

